# Prevalence, awareness, treatment, and control of hypertension in Bangladesh: Findings from National Demographic and Health Survey, 2017-18

**DOI:** 10.1101/2021.04.09.21255237

**Authors:** Md. Nuruzzaman Khan, John C. Oldroyd, Enayet K. Chowdhury, Mohammad Bellal Hossain, Juwel Rana, Stefano Renzetti, Rakibul M. Islam

## Abstract

**Objective:** To estimate the age-standardised prevalence, awareness, treatment, and control of hypertension and to identify their risk factors in Bangladeshi adults using the Bangladesh Demographic and Health Survey (BDHS) 2017-18 data.

**Methods:** Data from 12, 904 adults aged 18-95 years old, median (IQR) = 36 years (26-50), available from the most recent nationally representative BDHS 2017-18 were used. Hypertension was defined as having systolic blood pressure ≥140 mmHg and/or a diastolic blood pressure ≥90 mmHg, and/or taking anti-hypertensive drugs to control blood pressure. Age-standardised prevalence of hypertension and management were estimated with direct standardisation. A multilevel mixed-effects Poisson regression model with a robust variance was used to identify risk factors associated with hypertension and its awareness, treatment, and control.

**Results:** The overall age-standardised prevalence of hypertension was 26.2% (95% CI, 25.5-26.9); (men: 23.5%, women: 28.9%). Among those with hypertension (n=3531), 36.7% were aware that they had the condition, and only 31.1% received anti-hypertensive medication. Among those treated for hypertension (n=1306), 43.6% had controlled hypertension. Factors independently associated with hypertension were increasing age, higher body mass index, being women, having diabetes, and residing in selected administrative divisions. A declining trend of hypertension control was observed with increasing age and low education.

**Conclusions:** Hypertension is highly prevalent (1 in 4) in Bangladeshi adults, while awareness, treatment, and control are low. Irrespective of the risks associated with hypertension and its management, awareness, treatment, and control programmes should be given high priority in reducing hypertension and improving hypertension control in Bangladesh.

## 1. Introduction

Hypertension is one of the leading preventable causes of premature death and disability [1]. Globally, it accounted for ∼10.4 million deaths and 218 million disability-adjusted life-years in 2017 [2]. Hypertension is also a strong predictor of cardiovascular disease (CVD) and stroke [3]. Each 10mmHg rise in systolic blood pressure is associated with a 25% and 15% increased risk of CVD for women and men, respectively, [4] with similar risks for CVD mortality [4]. The lifetime risk of CVD at age 30 years is 63.3% in people with hypertension compared with 46.1% in people without hypertension [5]. People with hypertension develop CVD on average five years earlier than those without hypertension [5]. Economically, compared to those without hypertension, people with hypertension have substantially higher hospitalisation costs, outpatient care, and prescription medication [6].

In 2019, approximately 1.13 billion people had hypertension globally [1], compared with 972 million in 2000 [7]. Most people with hypertension are from low- and middle-income countries (LMICs) (1.04 billion) [8]. Over the past four decades, trends analysis shows a shift in the burden of hypertension from high-income countries (HICs) to LMICs, mostly in South Asia and sub-Saharan Africa [9]. From 2000 to 2010, the prevalence of awareness, treatment, and control increased far more in HICs, than LMICs [8]. This shift is important as hypertension awareness, treatment, and control indicate hypertension management and the associated burden of cardiovascular complications [10].

Bangladesh has experienced a rapid increase in hypertension [11]. A recent meta-analysis of 53 studies found that the overall prevalence of hypertension in Bangladeshi adults was 20%, varying from 1.10% to 75% [12]. The reasons for the variation in prevalence estimates are small sample size, use of data that are not nationally representative, and/or reporting of unstandardised estimates. A nationally representative study included in this meta-analysis reported that the age-standardised prevalence of hypertension was 27.1% in 2011 [13]. However, this study was limited to respondents aged 35 years and above; therefore, it cannot reflect the present situation where non-communicable diseases and their risk factors, including hypertension, are increasingly concentrated in the younger people [14]. Like hypertension, the management of hypertension is also poorly understood due to the small sample size and lack of nationally representative data [14,15]. Hence, understanding the prevalence and factors associated with hypertension and its management is decisive for improving treatment and control in Bangladesh.

To our knowledge, the age-standardised hypertension prevalence, awareness, treatment, control, and factors associated with these, among Bangladeshi adults 18 years and older using the latest Bangladesh Demographic and Health Survey (BDHS) 2017-18 data have not yet been estimated. Therefore, we aim to estimate the prevalence of hypertension and management and identify their risk factors in the Bangladeshi adult population 18 years and older. Results are examined in detail according to the individual, household, and community-level characteristics.

## 2. Methods

### 2.1 Study design and sample

Data for this study were extracted from the most recent nationally representative BDHS conducted between October 24, 2017 and March 15, 2018[11]. The BDHS is conducted every three years using two-stage stratified random sampling as part of the Demographic and Health Survey Program. In the first stage, 675 Primary Sampling Units (PSUs) were randomly selected from 293,579 PSUs. A total of 672 PSUs were included (urban: 192, rural: 480). The remaining three PSUs were not sampled due to flooding. In the second stage, 20,160 households were selected for data collection with 30 households from each selected PSU. Of them, interviews were completed in 19,457 households with a 96.5% inclusion rate. The BDHS 2017-18 collected blood pressure (BP) measurements from one-fourth of the selected households (7 to 8 HHs per cluster), which generated 4,864 households. There were 14,704 respondents aged 18 years and older in these selected households; 12,924 of them had BP measured with a response rate of 87.9% (Figure 1). The National Institute of Population Research and Training under the Ministry of Health and Family Welfare, Bangladesh, conducted this survey. Details of the sampling procedure have been published elsewhere [11].

**Figure 1:**
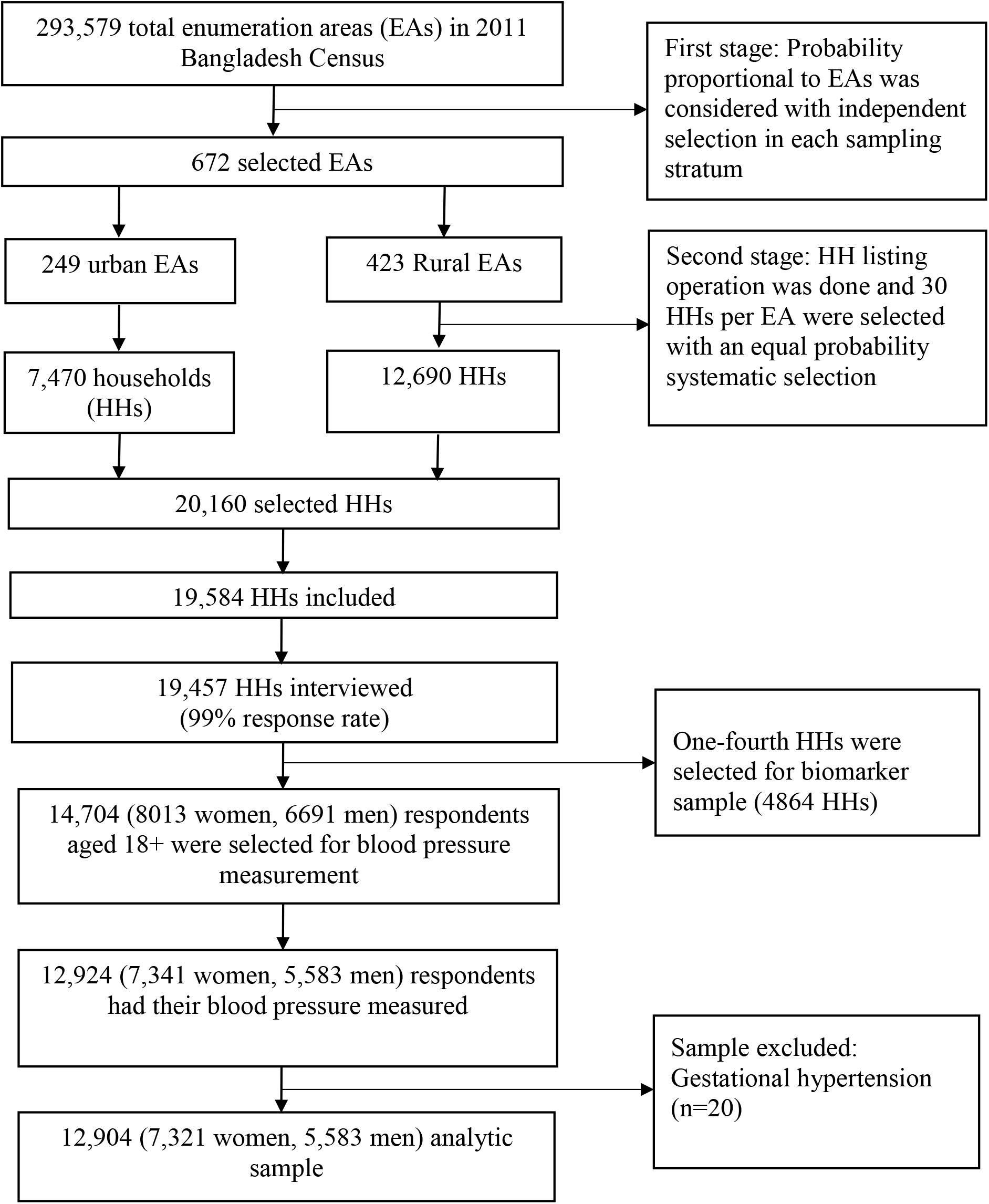
Flowchart of the study participants of the Bangladesh Demographic and Health Survey, 2017-18

### 2.2 Outcome measures

The prevalence, awareness, treatment, and control of hypertension were our outcomes of interest. The survey reported the systolic and diastolic BP measured in millimetres of mercury [mmHg]. Digital oscillometric BP measuring devices with automatic upper-arm inflation and automatic pressure release were used. Blood pressure was measured three times with an interval of at least 5 minutes, and the average of the second and third measurements was used as the individual BP. We classified an individual as hypertensive if he/she had (i) systolic blood pressure (SBP) ≥140 mmHg and/or a diastolic blood pressure (DBP) ≥90 mmHg, and/or (ii) taking any anti-hypertensive drugs to control BP. We used the BP cut-off value of 140/90 mmHg to define hypertension as per current National Guidelines for Management of Hypertension in Bangladesh [16], comparable with the 2018 European Society of Hypertension and European Society of Cardiology hypertension guidelines [17]. Awareness of hypertension was defined as hypertensive respondents, who identified based on the BP measurement, know their hypertensive status as a doctor or other health worker has told them. Treatment of hypertension was defined as self-reported use of prescribed antihypertensive medication to manage hypertension among those identified as hypertensive. Control of hypertension was defined as having an average SBP below 140mmHg and DBP below 90 mmHg among those were receiving antihypertensive medication.

### 2.3 Explanatory variables

We identified potential explanatory variables from previously published literature on hypertension in Bangladesh and other Southeast Asian countries [13,18,19]. The individual-level variables were age, sex, educational level, working status, body mass index (BMI), and diabetes status. Asian people are at greater risk of type 2 diabetes and cardiovascular disease at lower BMIs than the current WHO cut-off; thus, the BMI was categorised based on Asian cut-off (<18.5: Underweight; 18.5-23.0: Normal weight; 23.0-27.5: Overweight; ≥27.5: Obese) as suggested by the WHO expert consultation [20]. Diabetes was defined as a fasting blood glucose level greater than or equal to 7.0 mmol/L or self-reported use of glucose-lowering medication [21]. Wealth quintile was the household level variable based on household wealth index, which was constructed using principal component analysis from household’s durable and non-durable assets (e.g., televisions, bicycles, sources of drinking water, sanitation facilities, and construction materials of houses) [22] and expressed in five categories (poorest to richest). Community-level factors included were the place of residence (urban vs. rural) and region of residence (administrative divisions).

### 2.4 Statistical analysis

We used descriptive statistics to describe the individual, household, and community-level characteristics of the respondents. The prevalences of hypertension, awareness, treatment, and control were reported as crude and age-standardised estimates. The crude estimates were adjusted using a sampling weight and complex survey design to account for the data structure. The age distribution of the Bangladeshi population as reported in the 2011 Bangladesh national population and housing census was considered for the age-standardised prevalence. Variations between continuous and categorical variables were tested using the Mann-Whitney test and chi-square tests, respectively.

We used a multilevel mixed-effects Poisson regression model with a robust variance to identify factors associated with hypertension and its awareness, treatment, and control. The results were presented as a prevalence ratio (PR) with 95% confidence interval (95% CI). We used Poisson regression because the odds ratio estimated using logistic regression is usually overestimated if the outcome of interest is common, and the study design is cross-sectional [23]. Furthermore, in the BDHS, individuals were nested within the household; households were nested within the PSU/cluster. Therefore, our multilevel mixed-effects Poisson regression model accounts for these multiple hierarchies and dependency in data and the problem of overestimation. Three models were run separately for each outcome for different confounding factors at the individual, household, and community levels with progressive model-building technique. Model 1 was adjusted for only individual-level factors, Model 2 was adjusted for individual and household-level factors, and Model 3 was adjusted for individual, household, and community-level factors. Multi-collinearity and interaction between explanatory variables were tested before entering into models. All the statistical tests were two-sided, and a p-value <0.05 was considered statistically significant. The study was designed and reported following Strengthening the Reporting of Observational Studies in Epidemiology (STROBE) guidelines [24]. All analyses were performed using the statistical software package Stata (version 15.1; Stata Corp LP, College Station, Texas).

## 3. Results

### 3.1 Characteristics of the study sample

Of 12,924 participants who had their BP measured, 12,904 were included in this study (Figure 1). The total analytic sample by administrative divisions are shown in the map (Figure 2). Of these participants, 27% (n=3531) are classified as having hypertension. The individual, household, and community-level characteristics for the total respondents and their hypertension status are summarised in Table 1. The median (interquartile range, IQR) age of the study participants was 36 years (26-50), and 57% were female. A quarter of the participants had no formal education, 61% were employed, and 27% resided in urban areas. About 40% of participants were overweight/obese, and 13.9% had diabetes. People with hypertension were significantly different frompeople without hypertension across all characteristics (p = 0.014 to <0.001; Table 1).

**Figure 2:**
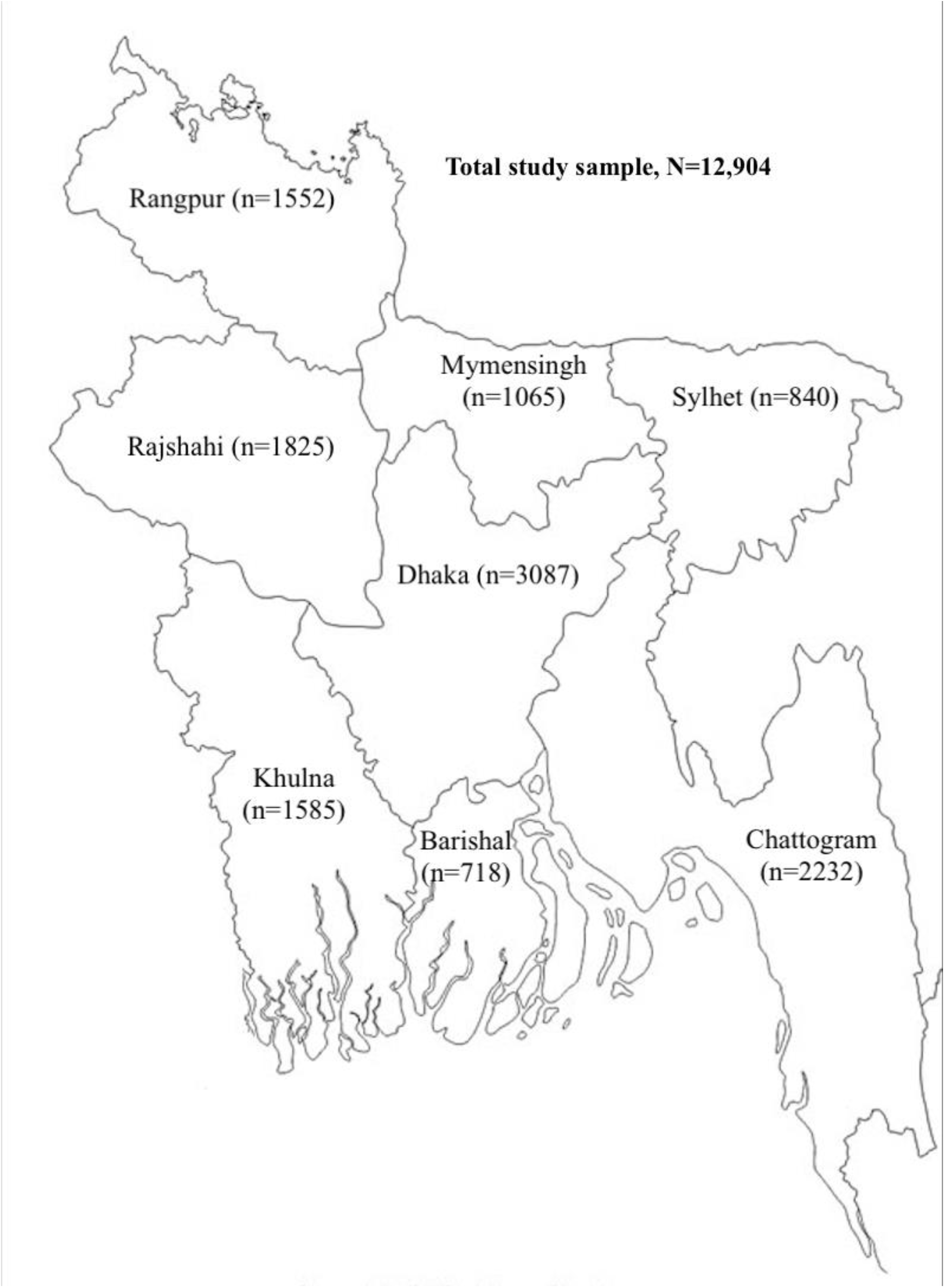
The administrative divisions of Bangladesh by the study sample size, BDHS 2017/18.

**Table 1:**
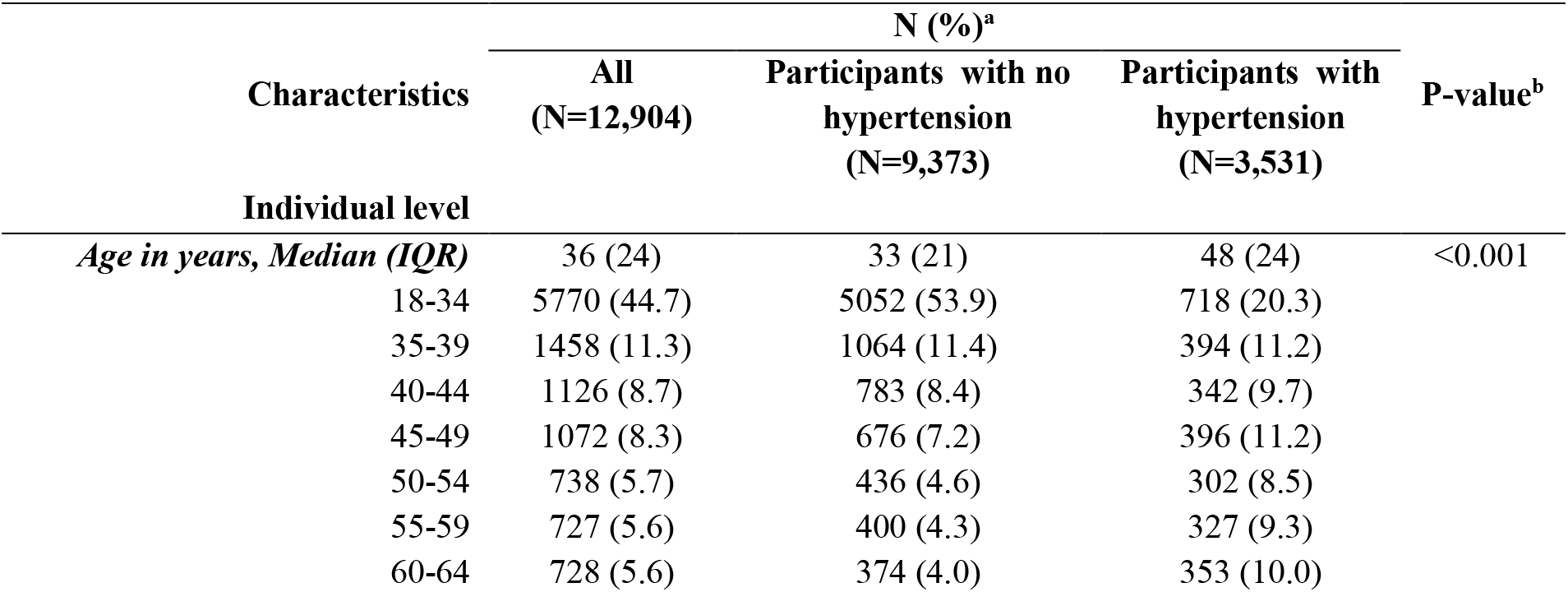
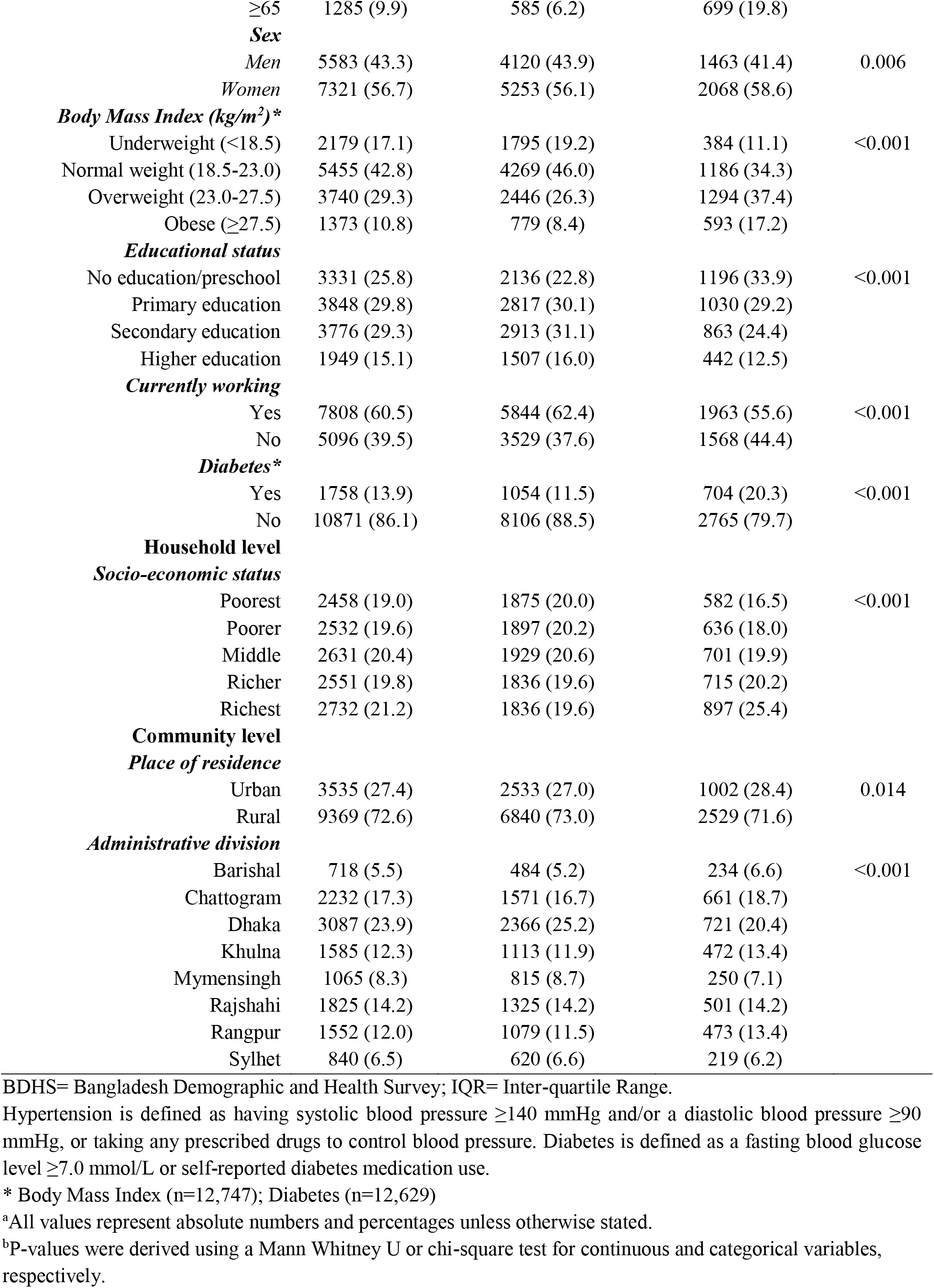
Characteristics of the study population

| Characteristics | N (%)<sup>a</sup> |  |  | P-value<sup>b</sup> |
| --- | --- | --- | --- | --- |
|  | All<br>(N=12,904) | Participants with no<br>hypertension<br>(N=9,373) | Participants with<br>hypertension<br>(N=3,531) |  |
| Individual level |  |  |  |  |
| *Age in years, Median (IQR)* | 36 (24) | 33 (21) | 48 (24) | <0.001 |
| 18-34 | 5770 (44.7) | 5052 (53.9) | 718 (20.3) |  |
| 35-39 | 1458 (11.3) | 1064 (11.4) | 394 (11.2) |  |
| 40-44 | 1126 (8.7) | 783 (8.4) | 342 (9.7) |  |
| 45-49 | 1072 (8.3) | 676 (7.2) | 396 (11.2) |  |
| 50-54 | 738 (5.7) | 436 (4.6) | 302 (8.5) |  |
| 55-59 | 727 (5.6) | 400 (4.3) | 327 (9.3) |  |
| 60-64 | 728 (5.6) | 374 (4.0) | 353 (10.0) |  |
| ≥65 | 1285 (9.9) | 585 (6.2) | 699 (19.8) |  |
| **Sex** |  |  |  |  |
| Men | 5583 (43.3) | 4120 (43.9) | 1463 (41.4) | 0.006 |
| Women | 7321 (56.7) | 5253 (56.1) | 2068 (58.6) |  |
| **Body Mass Index (kg/m<sup>2</sup>)\*** |  |  |  |  |
| Underweight (<18.5) | 2179 (17.1) | 1795 (19.2) | 384 (11.1) | <0.001 |
| Normal weight (18.5-23.0) | 5455 (42.8) | 4269 (46.0) | 1186 (34.3) |  |
| Overweight (23.0-27.5) | 3740 (29.3) | 2446 (26.3) | 1294 (37.4) |  |
| Obese (≥27.5) | 1373 (10.8) | 779 (8.4) | 593 (17.2) |  |
| **Educational status** |  |  |  |  |
| No education/preschool | 3331 (25.8) | 2136 (22.8) | 1196 (33.9) | <0.001 |
| Primary education | 3848 (29.8) | 2817 (30.1) | 1030 (29.2) |  |
| Secondary education | 3776 (29.3) | 2913 (31.1) | 863 (24.4) |  |
| Higher education | 1949 (15.1) | 1507 (16.0) | 442 (12.5) |  |
| **Currently working** |  |  |  |  |
| Yes | 7808 (60.5) | 5844 (62.4) | 1963 (55.6) | <0.001 |
| No | 5096 (39.5) | 3529 (37.6) | 1568 (44.4) |  |
| **Diabetes\*** |  |  |  |  |
| Yes | 1758 (13.9) | 1054 (11.5) | 704 (20.3) | <0.001 |
| No | 10871 (86.1) | 8106 (88.5) | 2765 (79.7) |  |
| **Household level** |  |  |  |  |
| **Socio-economic status** |  |  |  |  |
| Poorest | 2458 (19.0) | 1875 (20.0) | 582 (16.5) | <0.001 |
| Poorer | 2532 (19.6) | 1897 (20.2) | 636 (18.0) |  |
| Middle | 2631 (20.4) | 1929 (20.6) | 701 (19.9) |  |
| Richer | 2551 (19.8) | 1836 (19.6) | 715 (20.2) |  |
| Richest | 2732 (21.2) | 1836 (19.6) | 897 (25.4) |  |
| **Community level** |  |  |  |  |
| **Place of residence** |  |  |  |  |
| Urban | 3535 (27.4) | 2533 (27.0) | 1002 (28.4) | 0.014 |
| Rural | 9369 (72.6) | 6840 (73.0) | 2529 (71.6) |  |
| **Administrative division** |  |  |  |  |
| Barishal | 718 (5.5) | 484 (5.2) | 234 (6.6) | <0.001 |
| Chattogram | 2232 (17.3) | 1571 (16.7) | 661 (18.7) |  |
| Dhaka | 3087 (23.9) | 2366 (25.2) | 721 (20.4) |  |
| Khulna | 1585 (12.3) | 1113 (11.9) | 472 (13.4) |  |
| Mymensingh | 1065 (8.3) | 815 (8.7) | 250 (7.1) |  |
| Rajshahi | 1825 (14.2) | 1325 (14.2) | 501 (14.2) |  |
| Rangpur | 1552 (12.0) | 1079 (11.5) | 473 (13.4) |  |
| Sylhet | 840 (6.5) | 620 (6.6) | 219 (6.2) |  |

BDHS= Bangladesh Demographic and Health Survey; IQR= Inter-quartile Range.
Hypertension is defined as having systolic blood pressure ≥140 mmHg and/or a diastolic blood pressure ≥90 mmHg, or taking any prescribed drugs to control blood pressure. Diabetes is defined as a fasting blood glucose level ≥7.0 mmol/L or self-reported diabetes medication use.
\* Body Mass Index (n=12,747); Diabetes (n=12,629)
<sup>a</sup>All values represent absolute numbers and percentages unless otherwise stated.
<sup>b</sup>P-values were derived using a Mann Whitney U or chi-square test for continuous and categorical variables, respectively.

### 3.2 Prevalence of hypertension

The crude and age-standardised prevalence of hypertension by individual, household, and community-level factors are presented in Table 2. The overall age-standardized prevalence of hypertension was 26.2% (95% CI, 25.5-26.9); it was higher in women (28.9%, 95% CI, 27.9-29.9) than in men (23.5%, 95% CI, 22.5-24.6). The sex-specific age-standarised prevalence of hypertension is also higher in women than in men (Figure 3a). The prevalence of hypertension gradually increased by age and BMI; the highest prevalence was observed in people aged ≥65 years (56.4%, 95%CI 53.7-59.1) and obese (42.4%, 95%CI 40.0-44.8). The age-standardised prevalence of hypertension was also higher in people with diabetes (34.9%, 95%CI 32.7-37.1) than people without diabetes (25.0%, 95%CI 24.2-25.7).

**Table 2:** Crude and age standardized prevalence of hypertension, awareness, treatment and control in the Bangladeshi adults population, BDHS 2017-18

|  | Hypertension (3,531/12,904),<br>% (95% CI) |  | Awareness (1500/3531)<br>(% (95% CI) |  | Treatment (1306/3531)<br>% (95% CI) |  | Control (439/1306)<br>% (95% CI) |  |
| --- | --- | --- | --- | --- | --- | --- | --- | --- |
|  | Crude<br>prevalence | Standardized<br>prevalence* | Crude<br>prevalence | Standardized<br>prevalence* | Crude<br>prevalence | Standardized<br>prevalence* | Crude<br>prevalence | Standardized<br>prevalence* |
| **Overall** | 27.4 (26.3-28.4) | 26.2 (25.5-26.9) | 42.5 (40.5-44.5) | 36.7 (34.9-38.6) | 37.0 (35.2-38.9) | 31.1 (29.3-32.8) | 33.6 (30.7-36.7) | 43.6 (39.5-47.8) |
| **Age (years)** |  |  |  |  |  |  |  |  |
| 18-34 | 12.4 (11.4-13.5) | 12.8 (12.0-13.7) | 28.6 (25.1-32.4) | 27.5 (24.3-30.7) | 21.6 (18.5-25.1) | 21.3 (18.4-24.3) | 53.4 (44.3-62.3) | 53.5 (45.7-61.2) |
| 35-39 | 27.0 (24.6-29.5) | 27.0 (24.7-29.3) | 31.7 (26.8-37.1) | 34.5 (29.8-39.1) | 27.7 (23.0-32.8) | 30.0 (25.5-34.5) | 42.2 (32.4-52.7) | 40.5 (31.7-49.3) |
| 40-44 | 30.4 (27.5-33.5) | 31.5 (28.7-34.2) | 44.4 (38.7-50.2) | 44.1 (38.9-49.3) | 37.6 (31.9-43.6) | 38.5 (33.4-43.5) | 37.4 (28.3-47.5) | 39.4 (31.2-47.6) |
| 45-49 | 36.9 (33.5-40.6) | 37.6 (34.8-40.5) | 45.3 (39.9-50.8) | 46.9 (42.1-51.7) | 42.2 (36.8-47.8) | 43.0 (38.3-47.8) | 29.0 (22.1-36.9) | 29.1 (22.4-35.7) |
| 50-54 | 40.9 (36.9-44.8) | 41.5 (37.9-45.0) | 46.3 (40.6-52.2) | 49.2 (43.6-54.8) | 39.6 (34.1-45.4) | 42.4 (36.9-47.9) | 33.5 (24.8-43.4) | 34.9 (26.7-43.0) |
| 55-59 | 44.9 (41.0-49.0) | 46.5 (43.0-50.1) | 53.7 (47.9-59.3) | 54.9 (49.6-60.1) | 48.0 (42.3-53.7) | 49.4 (44.2-54.7) | 34.1 (26.6-42.4) | 31.8 (24.8-38.8) |
| 60-64 | 48.6 (44.6-52.5) | 49.1 (45.5-52.7) | 52.6 (47.2-57.9) | 52.5 (47.3-57.6) | 48.5 (43.2-53.8) | 48.1 (42.9-53.2) | 32.6 (25.1-41.1) | 31.4 (24.5-38.3) |
| ≥65 | 54.4 (51.5-57.4) | 56.4 (53.7-59.1) | 48.2 (44.4-52.1) | 50.4 (46.8-54.0) | 42.9 (39.1-46.8) | 44.9 (41.3-48.5) | 21.7 (17.5-26.5) | 25.4 (20.7-30.0) |
| **Sex** |  |  |  |  |  |  |  |  |
| Men | 26.2 (24.8-27.7) | 23.5 (22.5-24.6) | 33.4 (30.8-36.2) | 26.5 (23.8-29.1) | 28.7 (26.3-31.3) | 21.9 (19.5-24.2) | 31.7 (27.0-36.7) | 46.0 (37.7-54.3) |
| Women | 28.2 (27.1-29.4) | 28.9 (27.9-29.9) | 48.8 (46.3-51.4) | 43.5 (41.0-46.0) | 42.9 (40.5-45.3) | 37.2 (34.8-39.5) | 34.5 (31.0-38.3) | 42.9 (38.9-47.6) |
| **Body Mass Index (kg/m<sup>2</sup>)** |  |  |  |  |  |  |  |  |
| Underweight (<18.5) | 17.6 (15.9-19.4) | 14.4 (13.0-15.7) | 31.3 (26.7-36.3) | 26.6 (20.6-32.6) | 26.3 (22.0-31.1) | 22.6 (17.1-28.2) | 32.1 (23.6-42.0) | 41.7 (24.0-59.3) |
| Normal weight (18.5-23.0) | 21.7 (20.5-23.0) | 21.1 (20.1-22.1) | 39.9 (35.0-41.0) | 31.0 (27.8-34.1) | 32.3 (29.5-35.3) | 26.3 (23.3-29.3) | 35.0 (30.1-40.3) | 48.3 (40.3-56.2) |
| Overweight (23.0-27.5) | 34.6 (32.8-36.4) | 33.9 (32.5-35.3) | 45.0 (42.0-48.2) | 39.0 (36.0-41.9) | 40.2 (37.2-43.3) | 33.8 (31.1-36.6) | 29.8 (25.4-34.6) | 37.1 (30.9-43.3) |
| Obese (≥27.5) | 43.2 (40.6-45.9) | 42.4 (40.0-44.8) | 53.3 (49.0-57.6) | 49.5 (45.0-54.0) | 46.5 (42.3-50.8) | 40.9 (36.7-45.1) | 40.0 (34.0-46.2) | 50.0 (42.0-57.9) |
| **Educational status** |  |  |  |  |  |  |  |  |
| No education/preschool | 35.9 (33.9-37.9) | 23.5 (21.7-25.3) | 43.4 (40.1-46.8) | 31.2 (25.2-37.2) | 38.9 (35.7-42.3) | 26.4 (20.9-31.9) | 26.7 (22.5-31.4) | 36.1 (17.8-54.4) |
| Primary education | 26.8 (25.2-28.5) | 25.7 (24.4-27.0) | 44.2 (40.9-47.5) | 38.0 (34.5-41.6) | 37.8 (34.7-41.0) | 32.7 (29.3-36.1) | 35.6 (30.4-41.2) | 45.3 (37.9-52.7) |
| Secondary education | 22.8 (21.3-24.4) | 28.8 (27.3-30.3) | 41.6 (38.0-45.4) | 39.3 (36.0-42.6) | 36.6 (33.1-40.3) | 33.8 (30.7-36.9) | 37.9 (31.7-44.5) | 41.4 (34.8-48.0) |
| Higher education | 22.7 (20.6-24.9) | 30.7 (28.6-32.7) | 37.6 (33.2-42.1) | 37.0 (33.0-40.9) | 30.7 (26.7-35.1) | 31.0 (27.3-34.7) | 41.7 (33.4-50.4) | 51.0 (42.0-60.0) |
| **Currently working** |  |  |  |  |  |  |  |  |
| Yes | 25.1 (23.9-26.4) | 24.3 (23.3-25.2) | 37.0 (34.6-39.4) | 32.9 (30.5-35.3) | 31.6 (29.3-34.0) | 27.4 (25.1-29.6) | 34.4 (30.2-38.8) | 44.9 (39.3-50.5) |
| No | 30.7 (29.4-32.1) | 30.0 (28.7-31.2) | 49.4 (46.5-52.3) | 42.1 (39.1-45.0) | 43.8 (41.0-46.6) | 36.4 (33.6-39.2) | 32.9 (29.0-37.1) | 41.7 (35.7-47.8) |
| **Diabetes** |  |  |  |  |  |  |  |  |
| Yes | 40.0 (37.3-42.8) | 34.9 (32.7-37.1) | 54.2 (50.2-58.2) | 44.6 (39.7-49.5) | 49.4 (45.3-53.4) | 39.9 (35.2-44.6) | 30.3 (25.2-36.0) | 32.7 (23.9-41.5) |
| No | 25.4 (24.4-26.5) | 25.0 (24.2-25.7) | 39.6 (37.4-41.9) | 34.9 (32.8-36.9) | 34.1 (32.1-36.1) | 29.0 (27.1-30.9) | 34.6 (31.2-38.2) | 46.0 (41.4-50.7) |
| **Wealth quintile** |  |  |  |  |  |  |  |  |
| Lowest | 23.7 (21.8-25.7) | 20.7 (19.2-22.1) | 32.3 (28.0-36.9) | 25.8 (21.4-30.2) | 27.6 (23.9-31.6) | 20.9 (17.0-24.8) | 31.3 (23.5-40.3) | 42.8 (26.9-58.7) |
| Second | 25.1 (23.0-27.3) | 23.3 (21.7-24.9) | 36.5 (32.4-40.8) | 32.7 (28.4-37.0) | 30.3 (26.5-34.5) | 26.8 (22.8-30.8) | 32.8 (26.2-40.2) | 49.1 (39.3-58.9) |
| Middle | 26.6 (24.7-28.7) | 25.2 (23.6-26.8) | 42.0 (37.9-46.2) | 37.3 (32.8-41.7) | 37.4 (33.4-41.5)) | 32.5 (28.3 -36.8) | 31.9 (25.2-39.6) | 38.9 (30.0-46.9) |
| Fourth | 28.0 (25.9-30.3) | 28.0 (26.3-29.6) | 46.3 (42.2-50.4) | 39.7 (35.7-43.7) | 40.4 (36.5-44.3) | 33.1 (29.3-36.8) | 38.0 (31.9-44.6) | 38.5 (30.1-46.9) |
| Highest | 32.8 (30.9-34.8) | 32.7 (31.2-34.3) | 50.6 (46.9-54.4) | 42.4 (38.9-45.9) | 45.0 (41.5-48.5) | 36.9 (33.6-40.2) | 32.9 (28.2-37.9) | 47.8 (40.7-54.9) |
| *Place of residence* |  |  |  |  |  |  |  |  |
| Urban | 28.3 (26.6-30.1) | 28.7 (27.5-29.9) | 46.6 (43.3-50.0) | 39.3 (36.4-42.2) | 41.5 (38.4-44.7) | 34.8 (32.0-37.5) | 36.2 (31.5-41.2) | 44.1 (37.9-50.3) |
| Rural | 27.0 (25.8-28.2) | 24.9 (24.0-25.7) | 40.8 (38.4-43.3) | 35.2 (32.8-37.7) | 35.3 (33.0-37.6) | 28.8 (26.6-31.0) | 32.4 (28.8-36.3) | 43.2 (37.7-48.7) |
| *Administrative division* |  |  |  |  |  |  |  |  |
| Barishal | 32.6 (29.4-35.9) | 28.4 (26.2-30.6) | 44.3 (38.8-49.9) | 37.6 (31.7-43.4) | 38.9 (33.9-44.1) | 31.1 (25.8-36.5) | 34.5 (26.0-44.1) | 51.2 (38.6-63.7) |
| Chattogram | 29.6 (27.0-32.3) | 28.9 (26.9-30.9) | 44.8 (39.8-49.9) | 39.4 (34.7-44.0) | 41.3 (37.0-45.8) | 34.5 (30.1-38.9) | 41.7 (34.4-49.4) | 52.8 (43.5-62.1) |
| Dhaka | 23.3 (21.1-25.7) | 23.8 (21.9-25.6) | 45.2 (40.1-50.5) | 41.9 (36.3-47.5) | 40.7 (36.1-45.5) | 36.2 (30.8-41.5) | 32.6 (25.7-40.4) | 44.5 (34.0-55.0) |
| Khulna | 29.8 (27.1-32.7) | 26.8 (24.9-28.7) | 43.8 (38.6-49.2) | 39.3 (33.9-44.6) | 36.4 (31.1-42.0) | 32.2 (27.2-37.2) | 24.9 (19.5-31.3) | 32.1 (21.6-42.6) |
| Mymensingh | 23.5 (21.0-26.1) | 21.3 (19.4-23.3) | 42.5 (36.2-49.0) | 35.4 (29.5-41.3) | 39.2 (33.7-44.9) | 32.4 (26.7-38.0) | 48.7 (39.7-57.9) | 58.0 (44.2-71.8) |
| Rajshahi | 27.4 (24.7-30.3) | 25.9 (24.0-27.9) | 39.2 (34.2-44.5) | 29.5 (24.9-34.0) | 30.6 (26.0-35.7) | 21.9 (17.9-25.9) | 22.1 (15.8-29.9) | 32.0 (17.1-46.8) |
| Rangpur | 30.5 (28.3-32.7) | 28.9 (26.8-31.0) | 33.2 (28.6-38.2) | 28.8 (24.4-33.3) | 26.6 (22.4-31.2) | 21.6 (17.7-25.5) | 27.7 (20.5-36.3) | 25.3 (13.7-36.9) |
| Sylhet | 26.1 (23.4-29.0) | 25.3 (23.3-27.3) | 48.7 (42.2-55.3) | 44.0 (38.4-49.5) | 46.0 (39.5-52.7) | 41.4 (36.0-46.9) | 38.7 (30.3-47.8) | 48.2 (38.1-58.2) |

Data are % (95% CI). All CIs were logit transformed. \*Standardized to the 2011 Bangladesh Census population by age for the total population.
BDHS= Bangladesh Demographic and Health Survey.
Hypertension is defined as having systolic blood pressure $\geq 140$ mmHg and/or a diastolic blood pressure $\geq 90$ mmHg, or taking any prescribed drugs to control blood pressure. Awareness of hypertension was defined as a self-reported previous diagnosis of hypertension by a doctor or nurse in people with confirmed hypertension. Treatment of hypertension was defined as self-reported use of a prescription antihypertensive medication for management of hypertension. Control of hypertension was defined as receiving antihypertensive medication and having an average systolic blood pressure below 140mmHg and/or diastolic blood pressure below 90 mmHg.

**Figure 3:**
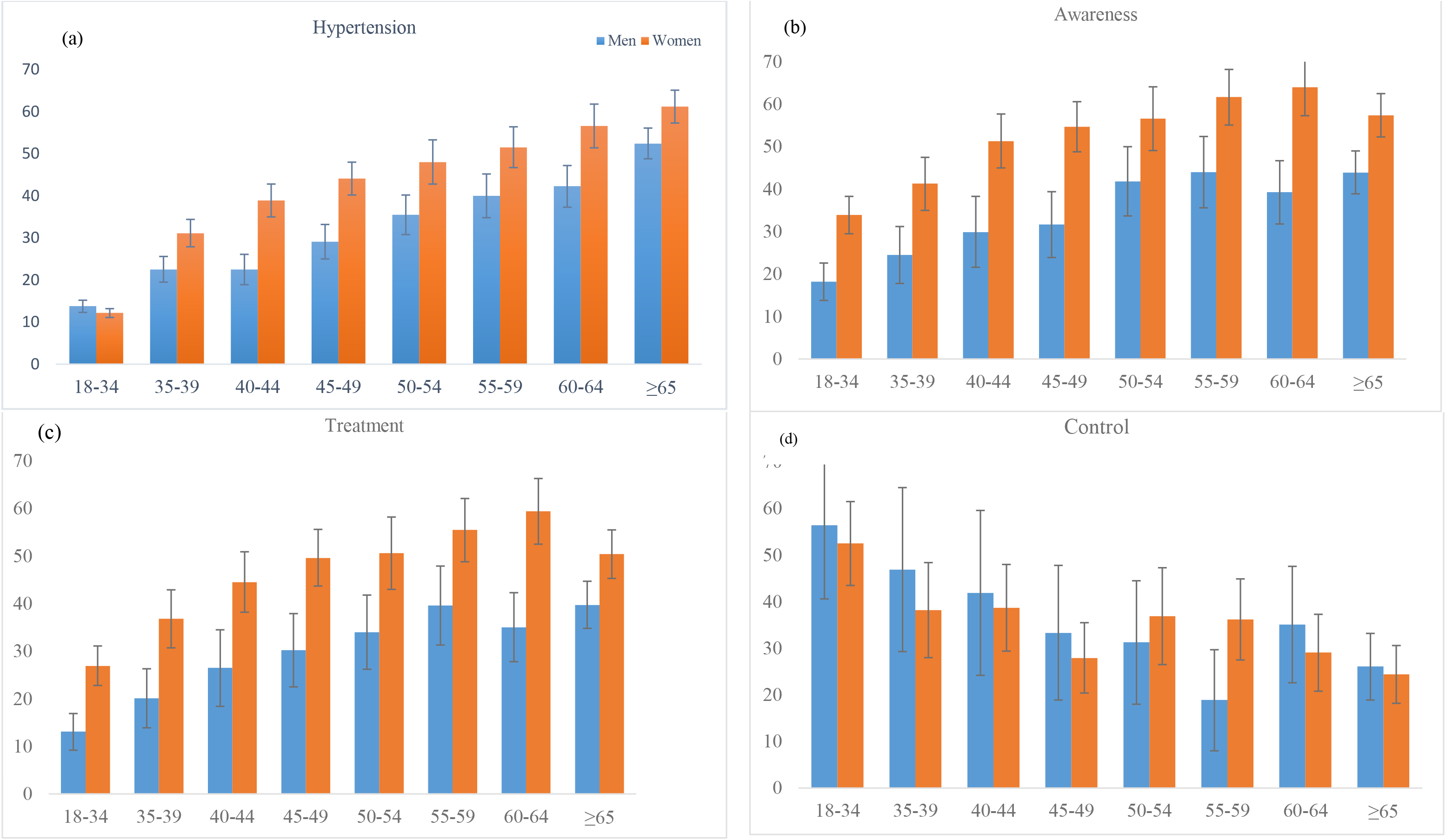
Age-standardized prevalence of (a) hypertension, (b) awareness, (c) treatment, and (d) control by age and sex in Bangladeshi adults

### 3.3 Awareness, treatment, and control of hypertension

Table 2 shows the crude and age-standardised prevalence of hypertension awareness, treatment, and control, by the individual, household, and community-level factors. In total, 43.5% (95%CI, 41.0-46.0) of hypertensive women and 26.5% (95%CI, 23.8-29.1) of hypertensive men were aware of having hypertension with an overall prevalence of 36.7% (95%CI, 34.9-38.6).

The sex-specific age-standarised prevalence of awareness was also higher in women than in men (Figure 3b). The proportion of awareness increased with increasing age up to 59 years, after which it declined; awareness also increased with BMI and wealth quintile. Among people who were hypertensive, only 31.1% (95%CI, 29.3-32.8) were on treatment, higher in women (37.2%, 95%CI, 34.8-39.5) than men (21.9%, 95% CI, 19.5-24.2). Among those treated for hypertension, 43.6% (95% CI, 39.5-47.8) had controlled hypertension. This rate was 15.1% (95% CI, 13.2-17.4) (results not shown in the table) when the total number of hypertensive respondents was considered. Overall, the proportion of controlled hypertension decreased with age but increased with the level of education. The age-standarised prevalence of treatment and control also had similar patterns between sexes (Figure 3c, 3d).

### 3.4 Factors associated with hypertension, awareness, control, and treatment

Three multilevel mixed-effects Poisson regression models were run to identify factors associated with hypertension, awareness, treatment, and control. The results for Model 3 (final model), adjusted for individual, household, and community-level factors, are shown in Table 3.The results for all other models are shown in Supplementary Table 1. Analyses showed that hypertension was independently associated with increasing age and increasing BMI, with the highest likelihood in people aged 65 years and older (PR 4.99, 95%CI 4.46-5.59), and in people with obesity (PR 1.87, 95%CI 1.71-2.04) compared with people aged 18-34 years and being normal weight, respectively. Women were more likely to be hypertensive (PR 1.15, 95%CI 1.07-1.23) than men. People with diabetes were also more likely to be hypertensive (PR 1.19, 95%CI 1.11-1.28) than people without diabetes. People underweight or living in Dhaka and Mymensingh divisions were less likely to be hypertensive than people of normal weight and living in the Barishal division. Other factors, including the level of education, employment status, wealth quintile, and place of residence, were not associated with hypertension.

**Table 3:**
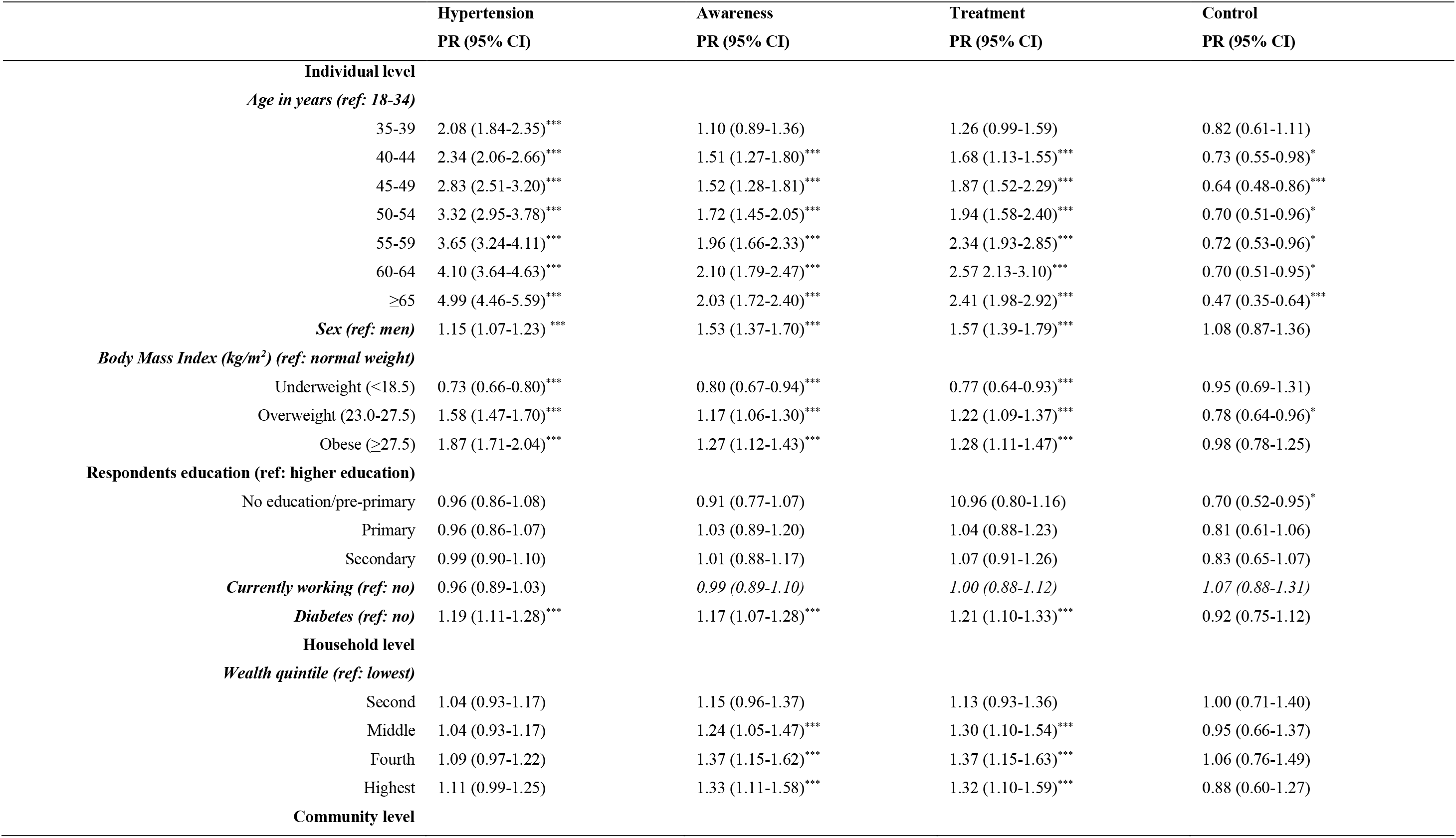
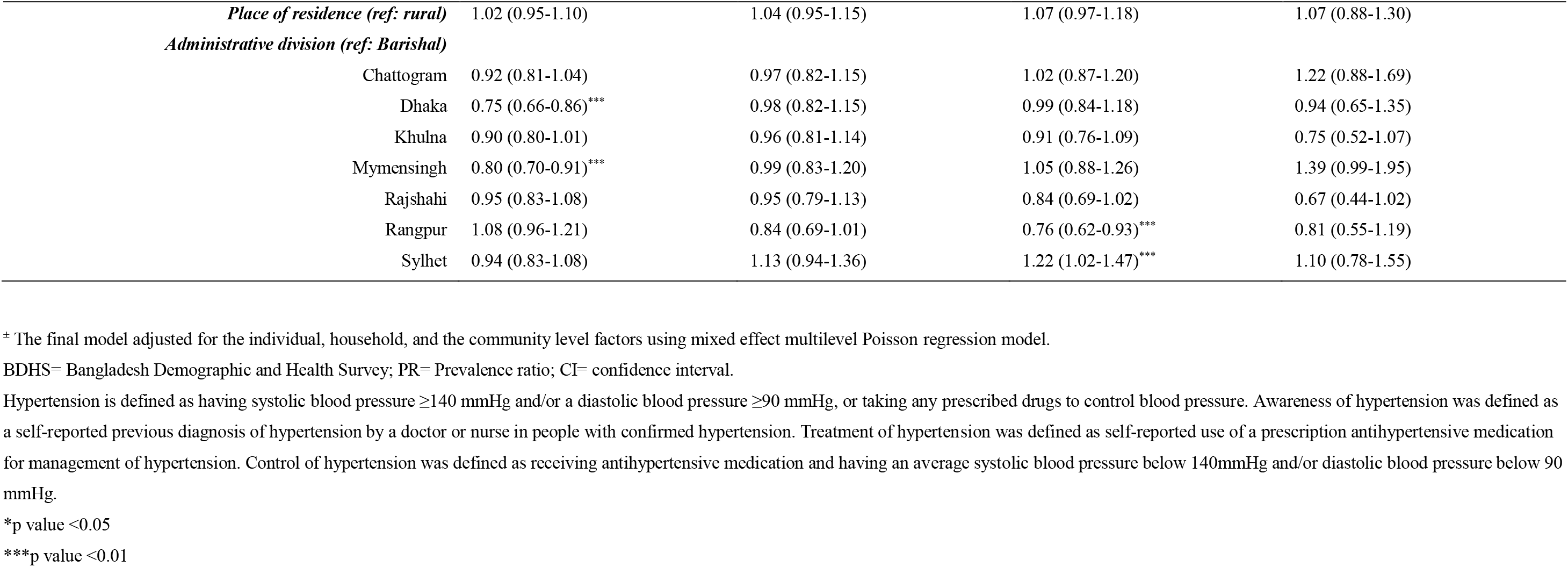
Factors associated with hypertension, awareness, treatment and control in adults aged ≥18 years in the Bangladeshi population, BDHS 2017-18

Awareness of hypertension was positively associated with age, sex, BMI, and wealth quintile. People with diabetes were also more likely to be aware (PR 1.17, 95%CI 1.07-1.28) compared with people without diabetes. Underweight people were less likely to be aware of hypertension compared with people of normal weight. Treatment of hypertension was positively associated with age, sex, BMI, diabetes status, wealth quintile, and administrative divisions but not with education, employment, or place of residence. Hypertension control was inversely associated with increasing age and not being educated.

## 4. Discussion

In this study in Bangladesh using the most recent nationally representative BDHS 2017-18 data, we estimated the age-standardised prevalence of hypertension, awareness, treatment, and control and identified risk factors associated with these conditions. Our findings showed that hypertension is highly prevalent in Bangladesh. Overall, one-fourth of survey adults aged 18 years and older had hypertension, accounting for 22.5 million adults in 2020. Among those with hypertension, over a third were aware of their condition, and only 31.1% were receiving treatment. Among those who received treatment, 43.6% had controlled hypertension.

Our study showed that the overall age-standardised prevalence of hypertension (26.2%) was higher than the overall age-standardised prevalence of South Asian countries (20.1%), but lower than overall LMICs (31.5%), and HICs (28.5%) [8]. The high prevalence of hypertension suggests that Bangladesh contributes significantly to the burden of hypertension in South-East Asia [25]. This is a challenge for the health system of Bangladesh, as the country has moved to the third phase of the epidemiological transition in terms of life expectancy at birth, with a double burden of disease. The rapid increment of hypertension in Bangladesh is most likely due to increasing prevalence and growth in the ageing population [8], urbanisation, and Westernised lifestyle, including excess dietary sodium intake and physical inactivity [8].

We found that the prevalence estimates of awareness, treatment, and control of hypertension were low. Our data suggest that Bangladesh has lower awareness and treatment and higher control of hypertension to other LMICs [8]. These findings highlight that although some progress has been made, greater treatment coverage is required, as treating less than a third of the hypertensive population is insufficient to achieve adequate BP control. The reasons for poor treatment could be explained by the lack of health literacy of hypertension and its consequenceses, care seeking behaviour or financial difficulties and/or no health insurance coverage that need be considered in future policies and programs [13]. Reasons for poor BP control in those treated for hypertension also need to be considered, including that hypertension occurs comorbidly with other risk factors among older people, which exacerbate each other to make BP control difficult.

Factors associated with hypertension were age, sex, BMI, diabetes, and selected administrative divisions of the country. Women had a higher prevalence of hypertension across all age categories than men, and hypertension awareness was also higher in women, consistent with previous studies in Bangladesh [13,18] and other countries [26]. We confirm obesity was associated with hypertension in this analysis, consistent with global literature [27,28]. Awareness and treatment were also significantly higher in people with obesity compared with normal weight. There was evidence in our study of more control of hypertension in people with overweight, not obesity, compared with normal weight. These findings are important as obesity is increasing in Bangladesh [29] due to the nutritional transition, characterised by the adoption of high-energy Westernised diets, and reduced physical activity [30].

In our study, the level of education and working status were not significantly associated with hypertension in the multivariate analysis though they were significant in the bivariate analysis. We found that the power of education and employment in explaining hypertension was diminished with the effect of BMI, as over 50% of the sample population who had higher education, were obese. Also, the observation that, ‘education is positively associated with working status’ is not true in the Bangladesh context, including our data. Evidence in Bangladesh shows that 37% women are working in paid employment, especially in garment sectors with no or primary education, while many women are highly educated but not engaged in paid employment (involved in unpaid household activities) [31]. Thus, the relationship between education and employment in Bangladesh is not linear, further explaining why hypertension was not associated with education or working status.

We also observed that awareness and treatment increased with ageing, whereas control decreased with age, in agreement with the current literature [26]. A possible explanation for the effects of advancing age is that individuals respond to health promotion (awareness) and have the financial resources to seek treatment as they grow older. However, by then, control is more difficult due to concurrent comorbidities. In addition,, poor medication adherence that arises due to the lack of health literacy, poor quality of drugs, or the use of traditional medicines, may also contribute to suboptimal BP control among those who are treated in Bangladesh [32]. Therefore, the educational and awareness-building interventions on the importance of monitoring and controlling BP should be considered in future policies and programmes.

We found that the likelihoods of hypertension awareness and treatment increased with increasing wealth quintiles for which there is inconclusive evidence in the literature [33,34]; however, we did not find differences in hypertension and its control across wealth quintiles. It may suggest that a similar rising pattern of hypertension in all segments of population is observed regardless of their wealth quintiles; however, people with improved socio-economic status are more likely to be aware of their conditions and received treatment. However, people from higher socio-economic status are more likely to consume more energy, and involved in white-collar jobs and follow sedentary lifestyles, which may increase the risk of high BP [34]. In contrast, several studies, including meta-analyses, suggest a higher prevalence of hypertension in lower socio-economic groups in HICs due to their higher smoking rates, elevated BMI, and lack of exercise than higher socio-economic groups [33,35,36].

In line with existing literature [37], our analysis showed that the prevalence of hypertension, awareness, and treatment was higher in people with diabetes than people without diabetes. However, fewer people with diabetes achieved control of their hypertension compared to people without diabetes. Both hypertension and diabetes contribute to macro and microvascular complications accelerating cardiovascular disease. Antihypertensive treatment reduces macro- and microvascular complications in people with diabetes and forms part of the diabetes management. [38].

Hypertension is a modifiable risk factor, and dietary and pharmaceutical interventions effectively control BP [39,40]. Despite this evidence, only 13.8% of adults with hypertension and 37.1% of people with treated hypertension worldwide had their BP controlled in 2010 [8]. Compared to this, our results show that overall control was better in Bangladesh despite resource contraints. Furthermore, there is evidence that interventions in Bangladesh to raise awareness using simple health messages to people with hypertension reduces BP modestly and improves BP control [41]. These are important as even small reductions in BP may have clinically important effects on hypertension reduction at a population level and future CVD and associated mortality [42].

The policy implications of our findings are that the management of hypertension in Bangladesh will require concerted efforts in primary prevention to raise awareness, including interventions to increase physical activity, achieve a healthy weight, consume diets high in fruits, vegetables and low in saturated fat. Our results also suggest that more investment, particularly in obesity management, is crucial to achieving better control at a population level. Delaying the onset of complications should be given priority through the aggressive clinical management of both hypertension and diabetes. In addition to interventions in high-risk individuals, more national population targeted approaches are needed in Bangladesh, similar to the national salt reduction programme planned for India [43], the UK Food Standards Agency Salt Reduction Strategy [44], and the WHO SHAKE intervention for salt reduction [45]. Finally, barriers to hypertension control within the health care system in Bangladesh need to be addressed at a policy level, such as access to health services, shortage of medicine supplies, long travel distances, long waiting times, and high costs [46].

The strengths of the study are that it used a large, nationally representative dataset suggesting the findings have external validity. A further strength is that clinical variables, including BP and body weight, and height, were measured using high-quality techniques. Our multilevel mixed-effects Poisson regression corrects the overestimation of effects size produced by conventional logistic regression employed in cross-sectional studies, and increased the precision of the findings. However, this was a cross-sectional study, which limits our ability to infer causal relationships. Further limitations are that we define hypertension and BP control based on BP records at a single time point, and anti-hypertensive medication use was self-reported. Moreover, a significant proportion of people with hypertension were not asked about their treatment status as they were unaware of the condition. Besides, a significant portion of people uses non-prescription medicines to control BP; this data was not collected, therefore, not adjusted for in the analyses. Dietary, physical activity, and smoking data were not collected as such could not be controlled for in the analyses though they are important predictors of hypertension.

## 5. Conclusion

Hypertension is highly prevalent (1 in 4) in Bangladeshi adults, while awareness, treatment, and control are low. Therefore, irrespective of risks associated with hypertension and its management, awareness, and control programmes should be given high priority in reducing hypertension and improving hypertension control in Bangladesh.

## Data Availability

Data can be available on request

## 6. Abbreviations

CVD: Cardiovascular disease
LMICs: Low- and middle-income countries
HICs: High income countries
BDHS: Bangladesh Demographic and Health Survey
PSU: Primary sampling units
SBS: systolic blood pressure
DBP: diastolic blood pressure
WHO: World Health Organization
PR: Prevalence Ratio, 95%
CI: 95% Confidence Interval
BMI: Body Mass Index

## 8. Consent for publication

Not applicable

## 9. Ethics approval and consent to participate

The survey, including biomarker measurements, was approved by the institutional review board at ICF and the Bangladesh Medical Research Council. Informed consent was obtained from all participants.

## 11. Availability of data and materials

The datasets used and analyzed during the current study are available from the Measure DHs website: https://dhsprogram.com/data/available-datasets.cfm

## 12. Acknowledgement

The authors thank to MEASURE DHS for granting access to the BDHS 2017-18 data.

---

**Supplementary table 1:**
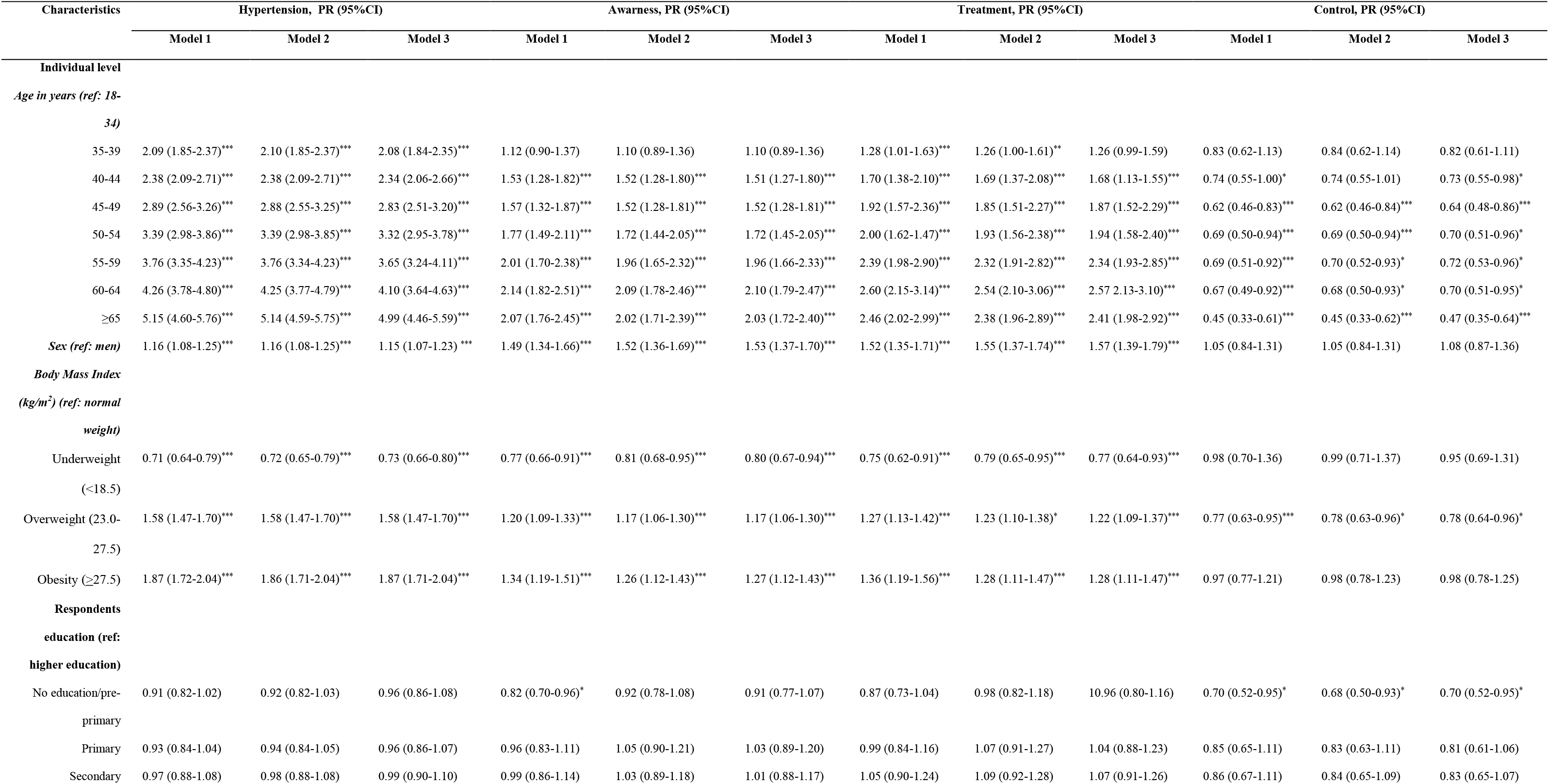
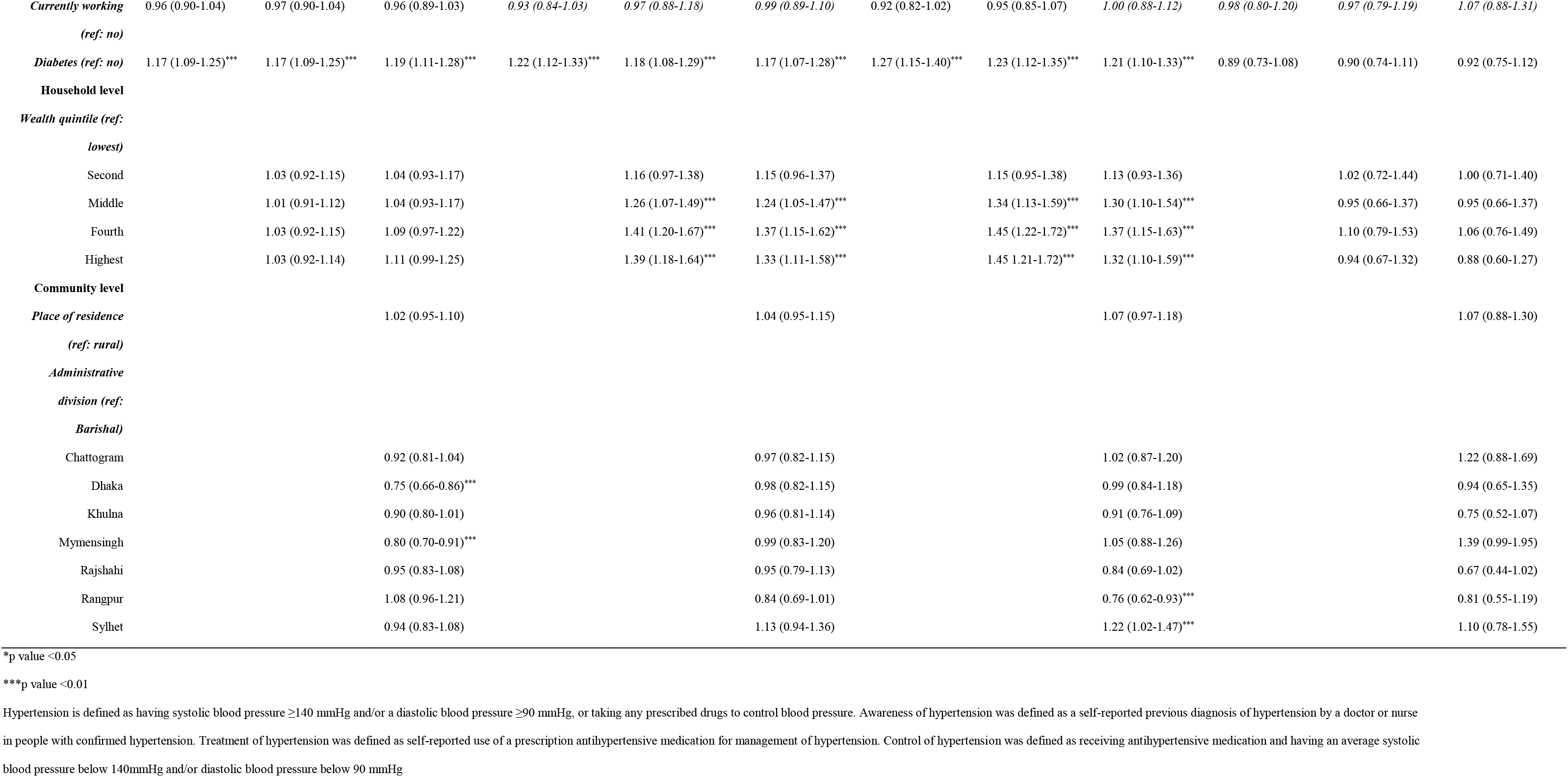
Factors associated with hypertension, awareness, treatment and control in adults aged ≥18 years in the Bangladeshi population, BDHS 2017-18

